# Mother-infant linked UK electronic birth cohorts representing 17.5 million births harmonised to the OMOP common data model

**DOI:** 10.64898/2026.03.23.26349078

**Authors:** M.J. Seaborne, S. Durbaba, A. Mendez-Villalon, T.C. Giles, A. González-Izquierdo, A. Hough, C. Sanchez-Soriano, N. Kennedy, H.E. Snell, N. Cockburn, L. Magee, K. Nirantharakumar, L. Poston, R.M. Reynolds, G. Santorelli, S. Brophy

## Abstract

We describe the harmonisation of five UK electronic birth cohorts to the Observational Medical Outcomes Partnership (OMOP) Common Data Model, creating a large-scale, standardised resource for maternal and child health research. The Mother and Infant Research Data Analysis (MIREDA) partnership developed and implemented reproducible guidelines for mapping maternal–infant relationships and identifying pregnancy episodes within routinely collected healthcare data. Cohorts from England, Scotland, and Wales were transformed despite substantial heterogeneity in data structure, coding systems, and variable definitions. The resulting harmonised resource preserves each cohort as an independent dataset while enabling federated analyses to be conducted across sites without the need to share individual-level data. Collectively, the cohorts capture over 17.5 million live births, providing sufficient scale to investigate rare exposures and outcomes, support trial emulation, and evaluate population-level policy impacts across the UK. This article details the transformation pipeline and provides reusable methods to support extension to additional cohorts and networks. The harmonised datasets enable interoperable, reproducible research and facilitate cross-national comparative studies in maternal and child health.

## Background and Summary

Birth and pregnancy cohorts are essential for studying early-life determinants of health and disease across the life course^1,2^. These cohorts provide detailed data on maternal characteristics, pregnancy complications and birth outcomes, and are often linked to longitudinal administrative records such as education, social care, and other public datasets^3–5^. Large birth cohorts have contributed substantially to understanding disease prevention and population health; however, many rely on time-limited participant recruitment, which can compromise representativeness and lead to datasets becoming outdated^6^. Advances in data science and the increasing availability of routinely collected electronic health records (EHRs)^7^ now enable the development of large-scale, anonymised and continuously updated electronic birth cohorts^8^.

Despite this potential, there is currently no unified structure for harmonising maternal and infant data across electronic cohorts. Within the UK, existing electronic birth cohorts differ in study design, coding systems and accessibility, creating barriers to cross-cohort and cross-nation analyses.

The Mother and Infant Research Electronic Data Analysis (MIREDA) partnership has undertaken the task of harmonising five UK birth cohorts^9^ into the Observational Medical Outcomes Partnership (OMOP) Common Data Model (CDM)^9–11^. The participating cohorts are Born in Wales^12^, Born in Scotland^13^, Born in Bradford^14–16^, Born in South London (eLIXIR)^17^, and maternity and infant data available through the Clinical Practice Research Datalink (CPRD)^18^. Together these cohorts provide population-scale coverage of births recorded across England, Scotland and Wales.

To achieve a standardised representation across cohorts, records were transformed into the OMOP CDM, an internationally adopted framework for structuring healthcare data. However, applying the CDM to maternal and infant health data presents specific challenges, as the model is primarily designed around individual patients, whereas pregnancy research requires explicit linkage between mothers and infants. We developed a transformation approach that enables representation of pregnancy episodes and core birth-related variables within the standard CDM structure, while preserving relationships between individuals. This approach supports secure, federated analysis by maintaining all data within their respective Trusted Research Environments (TREs).

Previous approaches to representing pregnancy data within the OMOP CDM have typically either attributed pregnancy information solely to the mother or relied on non-standard extensions to represent mother–infant relationships^19–21^. The former approach limits the ability to follow children longitudinally, while the latter reduces interoperability with the standard model. In this work, maternal and infant records are linked using mechanisms available within the standard CDM structure, enabling pregnancy episodes and mother– infant relationships to be represented while preserving individual identifiers for both individuals.

Together, the harmonised MIREDA cohorts represent over 17.5 million live birth records spanning England, Scotland and Wales, with linkage to contextual data including education, social care and mental health records. Harmonisation within the OMOP CDM enables analytical code to be developed once and executed across multiple cohorts using federated approaches, producing comparable analyses without transferring raw data between sites.

This data descriptor outlines the processes used to transform maternal and infant EHR data from participating cohorts into the OMOP CDM, including the mapping strategies required to represent pregnancy episodes and mother–infant relationships within the standard model. It describes the structure and content of the resulting harmonised dataset and provides reproducible methods to support extension to additional cohorts. This resource enables interoperable, federated analyses and supports cross-national and international research in maternal and child health.

## Methods

Representing pregnancy data within the OMOP CDM requires explicit linkage between maternal and infant records. To address this, mother–infant relationships were implemented using OMOP CDM person-level linkages and pregnancy episode definitions derived from source records, enabling both maternal and infant records to be represented as distinct individuals within the data model.

### Input Data

The harmonised dataset comprises five UK electronic birth cohorts. These cohorts include routinely collected maternal and infant health records linked with administrative datasets. These are summarised in Table 1.

**Table 1:** Summary of cohorts’ datasets in MIREDA.

| Cohort | Birth records | % lowest IMD quintile* | Dates covered | Linkage types |
| --- | --- | --- | --- | --- |
| CPRD Gold/Aurum | ~ 16.5M | 15.4% | 2000-present | Primary + secondary healthcare |
| Born in Wales | 774,335 | 25.1% | 2000-present | Primary + secondary healthcare, education, social care |
| Born in Scotland | 5880 | 11.8% | 2022-present | Maternity EHR + outcomes |
| Born in Bradford | 11,775 | 70.1% | 2016-present | Primary + secondary healthcare + education |
| Born in South London (eLIXIR) | 52,900 | 17.8% | 2018-2023 | Primary and Secondary Care health care |

\*Expected 20% of any cohort will be in lowest quintile.

#### Born in Wales (BiW)

This cohort includes routinely collected health, education, national census and social care datasets linked through the Secure Anonymised Information Linkage (SAIL) Databank^22^. The resource contains records for all births in Wales since 2000, with additional survey data from approximately 4,500 families recruited since 2020^12,23,24^.

#### Born in Scotland (BiS)

This pilot cohort links maternity electronic health records from the TRAK clinical system with maternal and child outcome data derived from national administrative datasets^13,25,26^.

#### Born in Bradford (BiBBS/BiB4ALL)

This cohort is embedded within maternity care records from Bradford Royal Infirmary and links routine health data with education and social care records. The cohort is managed by the Bradford Institute for Health Research^14–16,27^.

#### Born in South London (eLIXIR)

This dataset links maternity and neonatal records from BadgerNet and EPIC systems with primary care records, mental health data from the South London and Maudsley NHS Trust (SLaM), Hospital Episode Statistics and Health Visitor datasets^17,28,29^.

#### Clinical Practice Research Datalink (CPRD) England

Birth cohort data were derived from the Clinical Practice Research Datalink primary care databases, which are subdivided into CPRD Gold (Vision system) and CPRD Aurum (EMIS system). Pregnancy and birth records were identified through linkage to Hospital Episode Statistics and the CPRD Pregnancy Register^18,29,30^ Each cohort operates within its own secure Trusted Research Environment (TRE) under local governance frameworks described previously^12–18^. Data access and linkage procedures are conducted under the oversight of local data controllers and employ pseudonymisation and encryption to protect patient confidentiality.

### Data Harmonisation Process – The Standard OMOP process

Data harmonisation was conducted using version v5.4 of the OMOP CDM^10,11,31^. All data transformations took place locally in their designated TREs, and the final datasets were saved according to OMOP CDM standards. A schematic overview of the harmonisation workflow is shown in Figure 1. The use of a common data model enables analytical code to be executed in a federated manner across cohorts, allowing results to be generated locally within each TRE and combined without sharing individual-level data.

**Figure 1.**
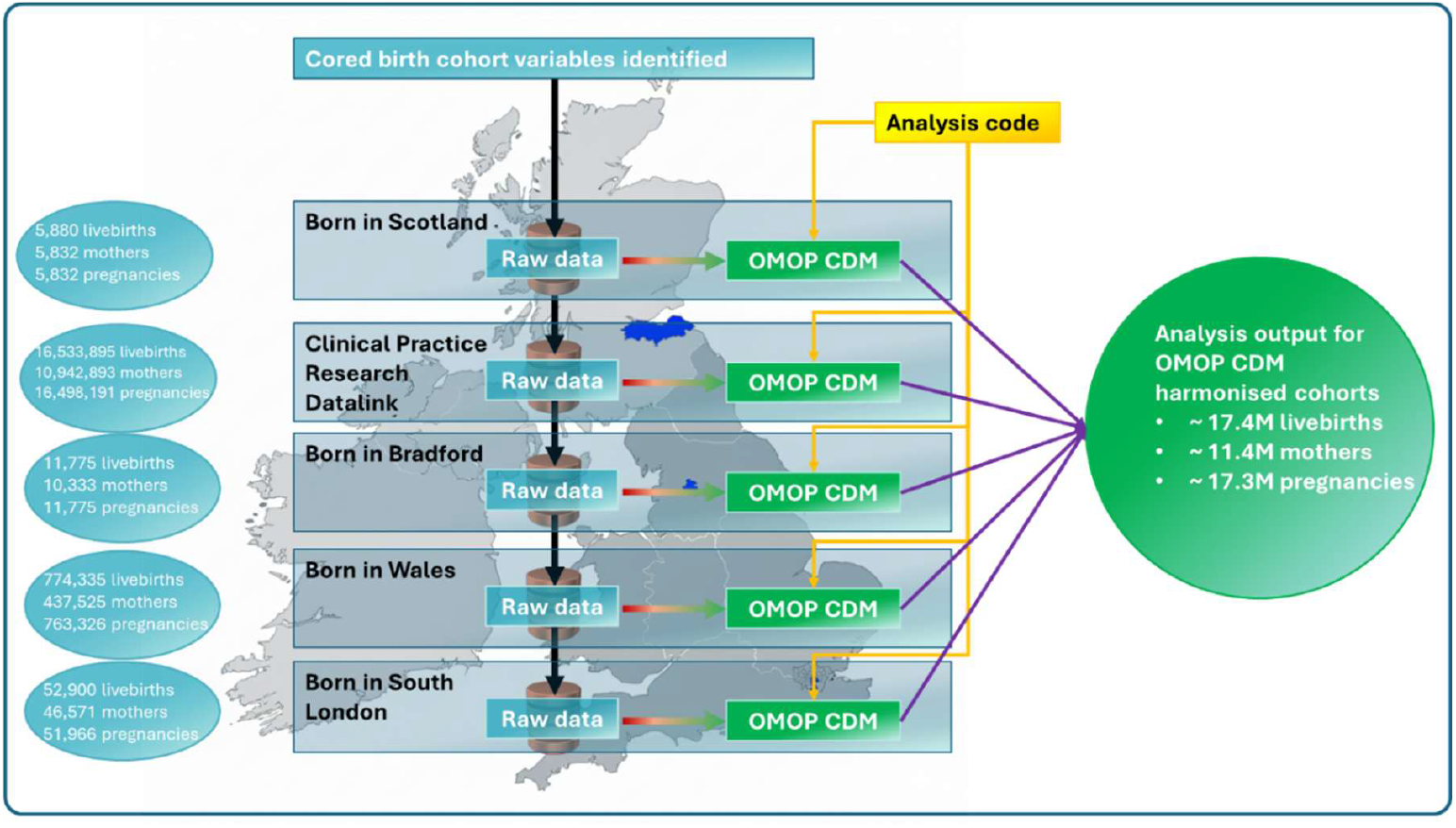
Overview process for harmonisation of UK birth cohorts into the OMOP CDM.

Core variables for harmonisation were defined through consensus within the MIREDA partnership prior to data extraction. These variables represent key maternal demographics, pregnancy characteristics, delivery information, and infant outcomes required to support longitudinal mother–infant analyses and are listed in Appendix A of the supplementary material.

### Extract, Transform, Load (ETL) Process

The harmonisation workflow followed a standard extract–transform–load (ETL) process implemented using tools developed by the Observational Health Data Sciences and Informatics (OHDSI) community.

#### Metadata profiling

Source database structures and vocabularies were profiled using WhiteRabbit^32^, which generates automated scan reports describing tables, variables and coding systems present within each dataset.

#### Vocabulary mapping

Scan reports were imported into Carrot Mapper^33–35^ to identify variables and values requiring mapping to OMOP CDM concepts. Mapping was conducted locally by analysts within each cohort environment.

Variables using standard clinical vocabularies supported by OHDSI (for example SNOMED CT, ICD-10 and OPCS-4) were automatically mapped where possible. Source variables using non-standard vocabularies were manually mapped to appropriate OMOP concepts by analysts using Carrot Mapper. Mapping rules are available in the MIREDA GitHub repositories^36^.

#### Transformation rule generation

Mapping decisions were exported as JSON transformation files describing the mapping rules required to convert source data to OMOP CDM tables.

#### Data transformation and loading

The transformation files were executed using Carrot Transform^37,38^, which was installed within each TRE to apply the ETL pipeline locally and load data into OMOP CDM format.

#### Cross-cohort harmonisation review

Mapping decisions and transformation rules were reviewed collaboratively across the MIREDA partnership to ensure consistent implementation across cohorts. The iterative workflow used to refine mappings and transformations is illustrated in Figure 2. The harmonisation pipeline was designed to be reproducible across sites, with mapping specifications and transformation rules defined in a standardised format to enable reuse in other OMOP CDM implementations.

**Figure 2:**
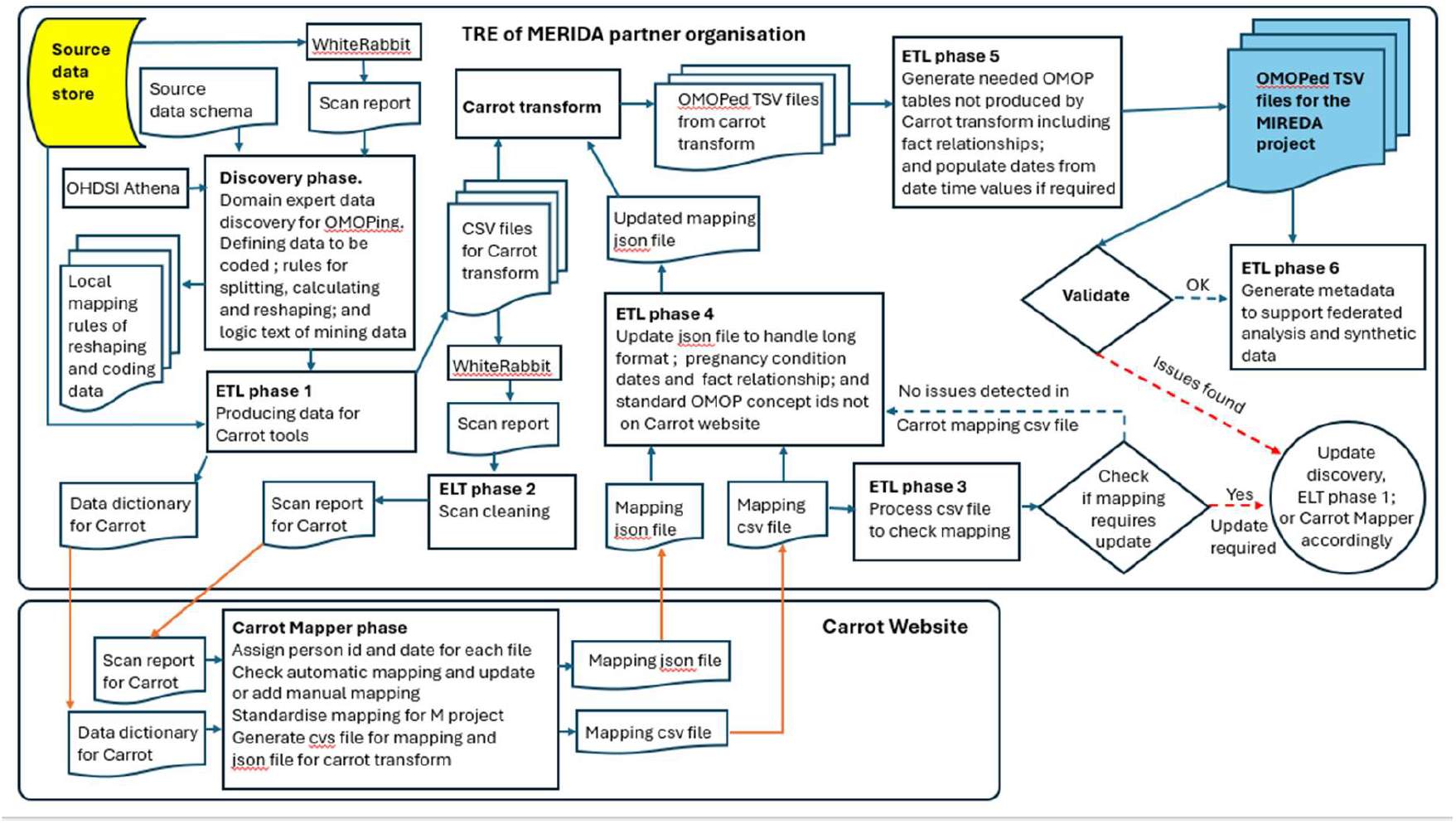
Data Harmonisation Process.

### Challenges and Solutions

#### Challenge 1: Mother-child linkage

The OMOP CDM is a person-centric data model and does not provide a standardised structure for representing relationships between maternal, pregnancy and infant records.

#### Solution

Mother-infant relationships were implemented using the fact_relationship table to link maternal and infant person identifiers, using OMOP relationship_concept_id for ‘mother’ (4248584) and ‘child’ (4285883).

Sibling relationships were also represented using the fact_relationship domain linking person identifiers with relationship_concept_id 4013484 for a twin sibling and 4292398 for a sibling from another pregnancy.

Pregnancy episodes were represented as condition_occurrence records using the condition_concept_id for pregnancy (4299535), with start and end dates derived from last menstrual period and delivery date. Infants from a pregnancy were linked using the relationship_concept_id 430541 (infant).

This solution implements existing mechanisms within standard OMOP CDM structures to form linkages without having to produce additional, non-standard domains. An alternative method used by others was to create non-standard OMOP perinatal expansion tables^21,39^. We have replicated this in parallel with our own method to facilitate possible future collaborations with these groups. See Figure *3*.

**Figure 3:**
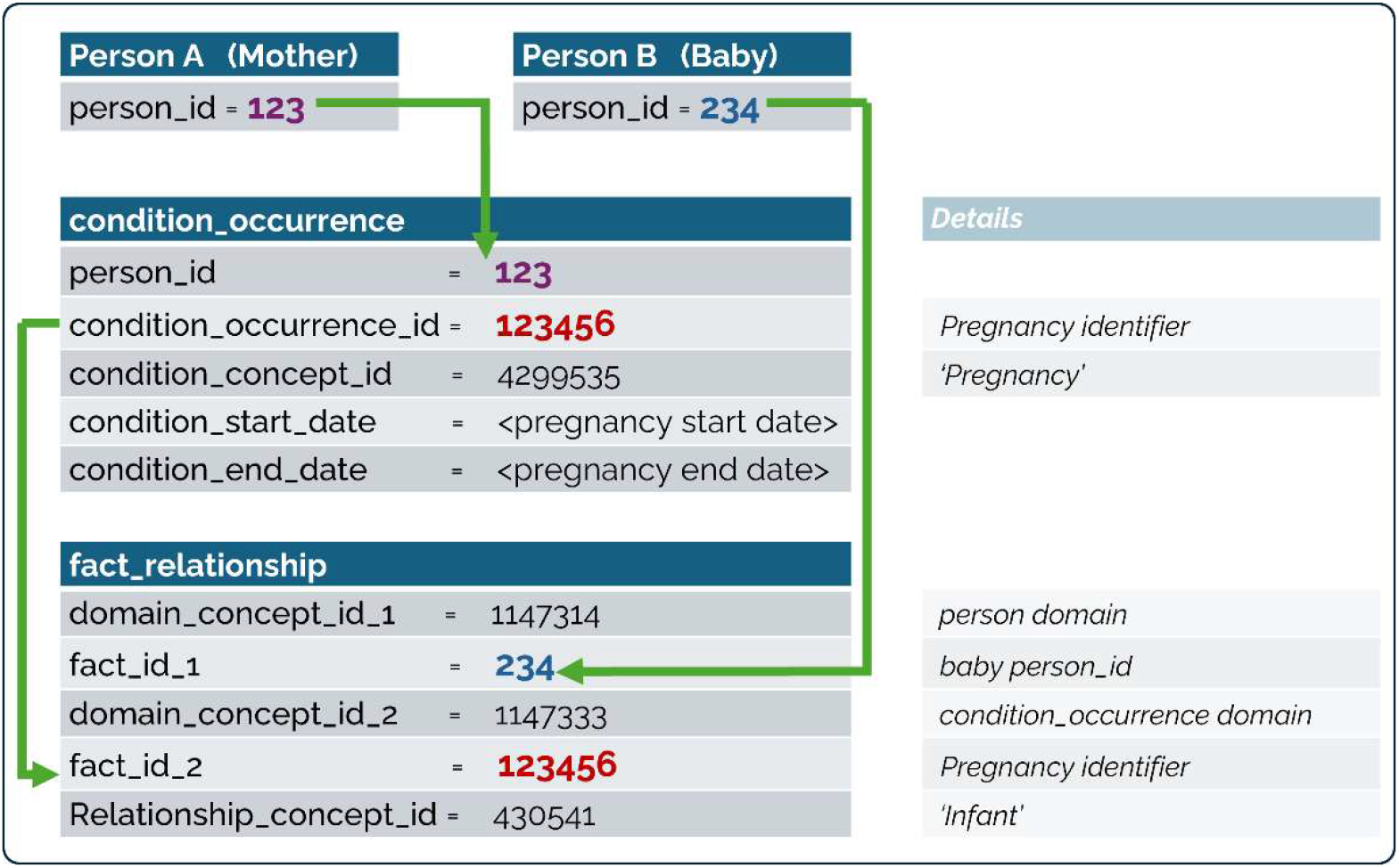
Linkage of mother, baby, and pregnancy

#### Challenge 2: Many-to-one mapping

Source variables may correspond to multiple valid OMOP concepts across different domains, leading to inconsistent mappings between cohorts.

#### Solution

A co-ordinated mapping strategy was implemented across all cohorts to ensure consistent assignment of variables to OMOP domains and concepts. For example, variables such as blood pressure and smoking status were standardised to consistent domain representations prior to transformation. Where necessary, source data were pre-processed to align with OMOP CDM structure (e.g. separation of systolic and diastolic measurements).

#### Challenge 3: White Rabbit formatting

WhiteRabbit scan reports present metadata in a long format that can separate related data variables, such as clinical codes and associated values, complicating interpretation of source data structure.

#### Solution

Where necessary, source data were restructured prior to mapping to preserve code-value relationships between variables. Although this required additional manual mapping pending future updates to Carrot Mapper, it enabled accurate transformation into OMOP CDM format.

#### Challenge 4: Location history

OMOP CDM stores only the most recent known address (location) for everyone, limiting the representation of time-varying location-based attributes.

#### Solution

A non-standard helper table (*‘location_history*’) was implemented, following the OMOP CDM v6.0 conventions^40^, to retain historical address records and support longitudinal analyses of time-varying demographic variables.

#### Challenge 5: Drug vocabulary

SNOMED and Read codes for drugs used in the UK are not fully aligned with OMOP standard vocabularies, which use RxNorm as its standard and ATC codes for classification.

#### Solution

Drug data were pre-processed using mapping tables to convert source codes to the UK DM+D standard, which has established mappings to RxNorm within the OMOP CDM. These mappings were then applied during transformation using Carrot Mapper. Additional reference tables were used to assign ATC classifications where required.

#### Challenge 6: OMOP CDM version updates

The OMOP CDM is updated biannually, requiring alignment across cohorts to maintain compatibility.

#### Solution

All cohorts synchronised updates to the OMOP CDM version used for transformation to ensure consistency of structure and vocabulary across sites.

#### Challenge 7: Granularity in ethnicity

Mapping UK ethnicity data to OMOP standard ‘race’ concepts results in a loss of detail due to differences between UK-specific classifications and OMOP standard vocabularies.

#### Solution

Broad ethnicity categories were mapped to standard OMOP ‘race’ concepts within the person table and constitute White (8527), Asian (8515), and Black (38003598), all others are considered missing. More detailed ethnicity information was preserved by mapping source categories to the observation domain using UK-specific classifications. This approach retains granularity while maintaining compatibility with standard OMOP analyses.

Note: Ethnicity mapping is still incomplete for all cohorts, but the mapping plan is available on the MIREDA GitHub.

### Limitation of using OMOP

The OMOP does not natively represent the absence of clinical conditions. As a result, source data indicating both the presence and confirmed absence of a condition (e.g. ‘has diabetes’ versus ‘no diabetes’) are reduced to representations of presence only. Individuals without a recorded condition may therefore represent either true absence or lack of recorded information. Similarly, source categories such as ‘unknown’ or ‘declined to answer’ are not consistently preserved within standard OMOP vocabularies, resulting in potential loss of detail for some variables.

More specific detail about these and other issues are available in the MIREDA GitHub repository^36^. In addition, where expected mappings were incorrect or errors arose in running the Carrot Transform software, R and python scripts were used to amend JSON files to affect the appropriate outcomes and shared between TREs where necessary and these files are available in the MIREDA GitHub.

### The Harmonised MIREDA datasets

The harmonised MIREDA datasets (see Table 2Table 5) comprise more than 17.5 million live births across England, Scotland and Wales. Approximately 99% of births are singletons, with multiple births (twins, triplets, etc) accounting for the remaining 1%. Term births account for 82% of records, with 7.1% preterm and 3.3% post-term deliveries. Mode of delivery was missing for 18.8% of births. Among records with available data, 25.4% were caesarean section deliveries.

**Table 2:** Breakdown of birth cohorts by births, mothers, and pregnancies.

|  | BiW | BiS | BiSL | BiB | CPRD Gold | CPRD AURUM | MIREDA |
| --- | --- | --- | --- | --- | --- | --- | --- |
| **All births, n** | 777,638 | 5,906 | 53,199 | 11,787 | 2,932,682 | 13,675,628 | 17,456,840 |
| **Live births, n (% births)** | 774,335 (99.6) | 5,880 (99.6) | 52,900 (99.4) | 11,775 (99.9) | 2,918,534 (99.5) | 13,615,361 (99.6) | 17,378,785 (99.6) |
| **Mothers, n\*** | 437,525 | 5,832 | 46,571 | 10,333 | 1,897,274 | 9,045,619 | 11,443,154 |
| **Pregnancies, n\*** | 763,326 | 5,832 | 51,966 | 11,775 | 2,912,775 | 13,585,416 | 17,331,090 |

\*For live births only.

**Table 3:** Labour and birth outcomes for live births only, where counts are the number of babies.

|  |  | BiW | BiS | BiSL | BiB | CPRD GOLD | CPRD AURUM | MIREDA |
| --- | --- | --- | --- | --- | --- | --- | --- | --- |
|  |  | n (%) | n (%) | n (%) | n (%) | n (%) | n (%) | n (%) |
| **Number of fetuses** | **Singleton** | 752,005 (97.1) | 5,788 (98.4) | 51,052 (96.5) | 9,043 (76.8) | 2,911,485 (99.8) | 13,476,181 (99) | 17,205,554 (99) |
|  | **Multiple** | 22,330 (2.9) | 92 (1.6) | 1,848 (3.5) | 2,732 (23.2) | 7,049 (0.2) | 139,180 (1) | 173,231 (1) |
| **Gestation** | **Very preterm (<28 weeks)** | 1,987 (0.3) | 17 (0.3) | 257 (0.5) | 11 (0.1) | 54,778 (1.9) | 237,823 (1.7) | 294,873 (1.7) |
|  | **Preterm (28-36 weeks)** | 54,287 (7) | 386 (6.6) | 3,779 (7.1) | 100 (0.8) | 165,682 (5.7) | 705,737 (5.2) | 929,971 (5.4) |
|  | **Term (37-41 week)** | 686,564 (88.7) | 5,375 (91.4) | 47,965 (90.7) | 1,654 (14) | 2,344,707 (80.3) | 11,193,294 (82.2) | 14,279,559 (82.2) |
|  | **Post term (42 weeks +)** | 31,497 (4.1) | 102 (1.7) | 899 (1.7) | 19 (0.2) | 118,975 (4.1) | 423,302 (3.1) | 574,794 (3.3) |
|  | **Not coded** | 0 (0) | 0 (0) | 0 (0) | 9,991 (84.8) | 234,392 (8) | 1,055,205 (7.8) | 1,299,588 (7.5) |
| **Low birth weight** | **Full term & weight <2500g** | 8,085 (1) | 153 (2.6) | 1,492 (2.8) | 0 (0) | 159,384 (5.5) | 794,447 (5.8) | 963,561 (5.5) |
| **Caesarean sections** | **C-section** | 154,087 (19.9) | 2,609 (44.4) | 21,027 (39.7) | 2,092 (17.8) | 587,041 (20.1) | 2,814,994 (20.7) | 3,581,850 (20.6) |
|  | **Non-C-section** | 393,390 (50.8) | 3,078 (52.3) | 31,675 (59.9) | 2,959 (25.1) | 1,744,101 (59.8) | 8,353,909 (61.4) | 10,529,112 (60.6) |
|  | **Not coded** | 226,858 (29.3) | 193 (3.3) | 198 (0.4) | 6,724 (57.1) | 587,392 (20.1) | 2,446,458 (18) | 3,267,823 (18.8) |
| **Birth mode** | **Elective C-section** | 69,108 (8.9) | 1,046 (17.8) | 9,090 (17.2) | 1,049 (8.9) | 264,108 (9) | 1,228,694 (9) | 1,573,095 (9.1) |
|  | **Emergency C-section** | 84,979 (11) | 1,563 (26.6) | 11,937 (22.6) | 1,043 (8.9) | 322,933 (11.1) | 1,586,300 (11.7) | 2,008,755 (11.6) |
|  | **Spontaneous** | 336,590 (43.5) | 2,363 (40.2) | 24,117 (45.6) | 2,605 (22.1) | 1,542,987 (52.9) | 7,382,271 (54.2) | 9,290,933 (53.5) |
|  | **Instrumental** | 56,800 (7.3) | 715 (12.2) | 7,558 (14.3) | 354 (3) | 157,355 (5.4) | 755,685 (5.6) | 978,467 (5.6) |
|  | **Not coded** | 226,858 (29.3) | 193 (3.3) | 198 (0.4) | 6,724 (57.1) | 631,151 (21.6) | 2,662,411 (19.6) | 3,527,535 (20.3) |

**Table 4:** Basic demographics of mothers in each cohort. All counts are for individual mothers.

|  |  | BiW | BiS | BiSL | BiB | CPRD GOLD | CPRD AURUM | MIREDA |
| --- | --- | --- | --- | --- | --- | --- | --- | --- |
|  |  | mean, [sd] | mean, [sd] | mean, [sd] | mean, [sd] | mean, [sd] | mean, [sd] | mean, [sd] |
|  | Age at delivery, years* | 29 [5.9] | 32 [5.21] | 33 [5.36] | 29 [5.72] | 29 [6.01] | 29 [5.97] | 29 [5.98] |
|  |  | n (%) | n (%) | n (%) | n (%) | n (%) | n (%) | n (%) |
| Ethnicity - raw data | White | 136,758 (31.3) | 4,539 (77.8) | 22,257 (47.8) | 3,399 (32.9) | 1,357,959 (71.6) | 6,230,144 (68.9) | 7,755,056 (44.7) |
|  | Black/Black British | 2,866 (0.7) | 281 (4.8) | 8,028 (17.2) | 158 (1.5) | 66,897 (3.5) | 506,754 (5.6) | 584,984 (3.4) |
|  | Asian/Asian British | 6,608 (1.5) | 538 (9.2) | 3,316 (7.1) | 3,152 (30.5) | 113,362 (6) | 834,279 (9.2) | 961,255 (5.5) |
|  | Mixed | 2,313 (0.5) | 87 (1.5) | 2,154 (4.6) | 131 (1.3) | 40,234 (2.1) | 252,949 (2.8) | 297,868 (1.7) |
|  | Other | 2,383 (0.5) | 137 (2.3) | 6,295 (13.5) | 334 (3.2) | 42,624 (2.2) | 263,655 (2.9) | 315,428 (1.8) |
|  | Missing | 286,597 (65.5) | 250 (4.3) | 4,521 (9.7) | 3,159 (30.6) | 276,198 (14.6) | 957,838 (10.6) | 1,528,563 (8.8) |

\*Where mothers had more than one birth, they were included in the mean age and standard deviation calculations for each birth.

**Table 5:** Maternal details during each pregnancy. Counts are numbers of pregnancies.

|  |  | BiW | BiS | BiSL | BiB | CPRD GOLD | CPRD AURUM | MIREDA |
| --- | --- | --- | --- | --- | --- | --- | --- | --- |
|  |  | n (%) | n (%) | n (%) | n (%) | n (%) | n (%) | n (%) |
| Age group | <20 years | 38,584 (5.1) | 101 (1.7) | 616 (1.2) | 463 (3.9) | 165,301 (5.7) | 709,949 (5.2) | 915,014 (5.3) |
|  | 20-29 years | 359,031 (47) | 1,635 (28) | 12,293 (23.7) | 5,826 (49.5) | 1,203,970 (41.3) | 5,509,710 (40.6) | 7,092,465 (40.9) |
|  | 30-39 years | 336,716 (44.1) | 3,784 (64.9) | 34,426 (66.2) | 5,043 (42.8) | 1,325,200 (45.5) | 6,154,159 (45.3) | 7,859,328 (45.3) |
|  | 40+ years | 28,995 (3.8) | 312 (5.3) | 4,631 (8.9) | 443 (3.8) | 218,304 (7.5) | 1,211,598 (8.9) | 1,464,283 (8.4) |
| Index of Multiple Deprivation (IMD), quintiles | 1 (most deprived) | 191,464 (25.1) | 686 (11.8) | 9,261 (17.8) | 8,258 (70.1) | 449,402 (15.4) | 2,113,828 (15.6) | 2,772,899 (16) |
|  | 2 | 160,604 (21) | 1,167 (20) | 19,140 (36.8) | 1,608 (13.7) | 577,948 (19.8) | 2,011,166 (14.8) | 2,771,633 (16) |
|  | 3 | 147,476 (19.3) | 924 (15.8) | 12,073 (23.2) | 601 (5.1) | 541,122 (18.6) | 2,667,176 (19.6) | 3,369,372 (19.4) |
|  | 4 | 132,212 (17.3) | 1,390 (23.8) | 6,427 (12.4) | 365 (3.1) | 638,609 (21.9) | 3,296,107 (24.3) | 4,075,110 (23.5) |
|  | 5 (least deprived) | 128,959 (16.9) | 1,464 (25.1) | 3,964 (7.6) | 143 (1.2) | 705,694 (24.2) | 3,497,139 (25.7) | 4,337,363 (25) |
|  | Not coded | 2,611 (0.3) | 201 (3.4) | 1,101 (2.1) | 800 (6.8) | 0 (0) | 0 (0) | 4,713 (0) |
| Behaviours | Smoking at booking visit | 31,328 (4.1) | 400 (6.9) | 1,628 (3.1) | 0 (0) | 73,236 (2.5) | 333,294 (2.5) | 439,886 (2.5) |
|  | Not coded | 731,998 (95.9) | 5,432 (93.1) | 50,338 (96.9) | 11,775 (100) | 2,839,539 (97.5) | 13,252,122 (97.5) | 16,891,204 (97.5) |
|  | Smoking at delivery | 44,646 (5.8) | 0 (0) | 3,064 (5.9) | 0 (0) | 12,340 (0.4) | 61,914 (0.5) | 121,964 (0.7) |
|  | Not coded | 718,680 (94.2) | 5,832 (100) | 48,902 (94.1) | 11,775 (100) | 2,900,435 (99.6) | 13,523,502 (99.5) | 17,209,126 (99.3) |
| Conditions arising during pregnancy | Diabetes (gestational) | 21,139 (2.8) | 0 (0) | 5,429 (10.4) | 0 (0) | 95,462 (3.3) | 486,395 (3.6) | 608,425 (3.5) |
|  | Not coded | 742,187 (97.2) | 5,832 (100) | 46,537 (89.6) | 11,775 (100) | 2,817,313 (96.7) | 13,099,021 (96.4) | 16,722,665 (96.5) |
|  | Hypertension* | 11,815 (1.5) | 0 (0) | 3,038 (5.8) | 63 (0.5) | 5,788 (0.2) | 27,610 (0.2) | 48,314 (0.3) |
|  | Not coded | 751,511 (98.5) | 5,832 (100) | 48,928 (94.2) | 11,712 (99.5) | 2,906,987 (99.8) | 13,557,806 (99.8) | 17,282,776 (99.7) |
|  | Pre-eclampsia/ eclampsia | 6,085 (0.8) | 0 (0) | 1,261 (2.4) | 34 (0.3) | 45,102 (1.5) | 215,780 (1.6) | 268,262 (1.5) |
|  | Not coded | 757,241 (99.2) | 5,832 (100) | 50,705 (97.6) | 11,741 (99.7) | 2,867,673 (98.5) | 13,369,636 (98.4) | 17,062,828 (98.5) |
|  | **Anaemia** | 9,104 (1.2) | 0 (0) | 1,346 (2.6) | 0 (0) | 145,964 (5) | 589,063 (4.3) | 745,477 (4.3) |
|  | **Not coded** | 754,222 (98.8) | 5,832 (100) | 50,620 (97.4) | 11,775 (100) | 2,766,811 (95) | 12,996,353 (95.7) | 16,585,613 (95.7) |
|  | **Depression** | 21,160 (2.8) | 428 (7.3) | 4,781 (9.2) | 0 (0) | 81,547 (2.8) | 298,769 (2.2) | 406,685 (2.3) |
|  | **Not coded** | 742,166 (97.2) | 5,404 (92.7) | 47,185 (90.8) | 11,775 (100) | 2,831,228 (97.2) | 13,286,647 (97.8) | 16,924,405 (97.7) |
| **Parity** | **Nulliparous** | 293,069 (38.4) | 2,206 (37.8) | 23,422 (45.1) | 891 (7.6) | 1,913,833 (65.7) | 9,108,847 (67) | 11,342,268 (65.4) |
|  | **Primiparous** | 204,581 (26.8) | 1,453 (24.9) | 13,701 (26.4) | 9,308 (79) | 702,779 (24.1) | 3,035,329 (22.3) | 3,967,151 (22.9) |
|  | **Multiparous** | 129,305 (16.9) | 661 (11.3) | 7,732 (14.9) | 132 (1.1) | 296,119 (10.2) | 1,440,929 (10.6) | 1,874,878 (10.8) |
|  | **Missing** | 136,371 (17.9) | 1,512 (25.9) | 7,111 (13.7) | 1,444 (12.3) | 44 (0) | 311 (0) | 146,793 (0.8) |

*\*Includes gestational hypertension and pre-eclampsia.*

The dataset includes 11,443,154 mothers, with a mean maternal age of 29.4 years at delivery. Smoking at pregnancy booking was recorded in 2.6% of mothers.

Significant variation in key variables is observed across cohorts, reflecting differences in population characteristics, data capture and regional healthcare systems. For example, elective caesarean section rates vary between 39.7% in London and 19.9% in Wales.

Post-term delivery rates vary from 3.1% in CPRD Aurum/4.1% in CPRD Gold, to 1.7% in London. These two findings suggest very different practices in different regions, the longer-term outcomes of which can be explored using these datasets.

File Structure and Formats.

Each cohort provides OMOP CDM-compliant tables within a relational database environment (PostgreSQL or equivalent). Core tables include:

- person - demographic data for mothers and infants.
- condition_occurrence - pregnancy episodes and diagnoses.
- measurement - clinical measurements (e.g., gestational age, birth weight).
- observation - clinical facts obtained through examination, questioning or procedures.
- drug_exposure - prescribed medications.
- procedure_occurrence – processes carried out with a diagnostic or therapeutic purpose.
- fact_relationship - links between mother, child, and pregnancy identifiers.
- Additional helper tables (e.g., location_history) retain information not natively supported within the OMOP CDM (e.g., historical address data).

Key variables include gestational age, mode of delivery, pregnancy identifiers (condition_occurrence_id) and relationship identifiers (relationship_concept_id). All concept IDs conform to the standard OMOP vocabulary definitions^41^.

The core OMOP CDM dataset can be linked to additional datasets, such as health visitor data, education data, pre- and post-pregnancy GP data, pre- and post-pregnancy hospital data, emergency department data, congenital abnormalities datasets and vaccination records. Where recognised UK coding is used for these datasets, cohorts have included them in addition to the core data as they were directly transformed using carrot tools. Range of content may vary in this regard.

### Technical Validation

Technical validation was conducted throughout the ETL process to ensure data quality and consistency. Compliance with the OMOP CDM version 5.4 specifications was verified using automated schema validation to confirm the presence and structure of required tables and fields. Concept mapping accuracy was assessed through manual review of a random sample of source codes against their corresponding OMOP standard concepts, including SNOMED CT, RxNorm, ICD-10 and OPCS4. Ambiguous and high-frequency codes were prioritised for review to minimise misclassification.

Mother–infant linkage was validated by comparing relationships recorded in the fact_relationship table with original linkage identifiers in the source datasets. Pregnancy episode integrity was assessed by comparing derived episode dates with gestational age measurements. Aggregate-level validation was conducted by comparing key indicators, such as caesarean section rates and preterm birth counts, with published cohort statistics to identify potential discrepancies arising from transformation. Harmonisation decisions were reviewed collaboratively across cohorts to ensure consistent implementation of mappings and transformations. These steps collectively ensured that the transformed data were accurate, internally consistent, and suitable for federated analysis.

### Data Usage

Upon successful application to use the data, researchers will work with our analysts to produce and run the code for analyses on the researchers’ behalf.

Key structural considerations include:

- Mother–infant relationships are defined within the fact_relationship table
- Pregnancy episodes can be identified through the condition_occurrence records and associated fact_relationship tables.
- Helper tables, including location_history, provide additional contextual information not natively supported by OMOP.
- Some concepts, such as the absence of disease, are not explicitly represented in OMOP and may appear as missing values.

Detailed mapping specifications and supporting documentation are available in the accompanying GitHub repository.

### Data Availability

The harmonised OMOP CDM datasets remain within the TREs of each contributing cohort and requests for access through the Health Data Research UK Gateway^42^. Analysis requires OMOP-compatible tools, such as OHDSI packages in R or Python.

### Code Availability

All scripts and specifications used to transform source data into OMOP CDM format are available in the MIREDA Partnership’s GitHub repositories. These include WhiteRabbit scan reports, Carrot Mapper^43^ mapping files, and documentation of manual transformation steps. WhiteRabbit, Carrot Mapper, and Carrot Transform are third-party packages; further information is available through the OHDSI and Carrot communities respectively. The OMOP CDM version implemented was 5.4, using the most recent vocabulary release. No proprietary code was used in this work. Researchers can reproduce the harmonisation process using these resources and the standard OHDSI toolset^44^.

### Resource Summary

The harmonised datasets provide a standardised representation of UK maternal and infant health records across multiple cohorts using the OMOP CDM. The resource supports large-scale, federated analyses and enables cross-cohort comparison while preserving data governance constraints.

## Conclusions

The harmonised UK dataset can be used to study rare conditions where large sample sizes are required, and to compare local practices, policies, and outcomes. However, it must be recognised that standardisation can result in data loss or reduced granularity. The OMOP CDM is designed for healthcare settings and does not cover non-health domains; therefore, linked data sources, such as education data, are not standardised within the OMOP CDM framework. Governance and access procedures differ between TREs, meaning that time is required to apply to each TRE for approval to access their OMOP data. Harmonisation must be iterative to support updates to both raw data and the OMOP model. This work presents a method for harmonising maternal and infant UK EHR data for federated analysis, and for standardising concepts to facilitate cross-nation collaborations in life course research.

### Future Development

Ongoing work within the MIREDA partnership includes completion of ethnicity harmonisation across the remaining cohorts. In parallel, development is underway to extend the data model by linking clinical events to additional CDM episode tables, enabling more explicit representation of pregnancy episodes and their associated events. At the time of this work, Northern Ireland datasets could not be harmonised due to system changes; efforts are currently in progress to obtain the necessary permissions to transform their CPRD data, with plans to harmonise data from the updated systems as they become available. Collaboration with Northern Ireland partners will include knowledge transfer from BiSL analysts who have transformed their data on the same system that Northern Ireland are implementing. In Scotland, the dataset is relatively recent and was initially based on a consented cohort, with many pregnancies not yet completed. Following the introduction of opt-out consent, substantially larger data volumes are expected, and future harmonisation will incorporate these expanded records. We will undertake pilot study for the use of a federated platform and develop this for future access to our cohorts. Finally, while each cohort currently uses country-specific indices of multiple deprivation, work is planned to develop a composite measure to support cross-cohort comparability.

## Data Availability

All data for is securely held at each cohort's Trusted Research Environment (TRE) and access is granted via the each TRE's request and governance frameworks.

https://github.com/MIREDA-Partnership

https://healthdatagateway.org/en/collection/119

## Author Contributions

MS, HS, and SB conceived the study design. MS, SD, DF, AH, and DM performed data extraction, transformation, and harmonisation. SB, KN, LP, and RR provided oversight and methodological guidance. All authors contributed to drafting and revising the manuscript and approved the final version.

## Acknowledgements

We thank Tisha Dasgupta at King’s College London for their contributions to data governance and patient and public involvement activities. We also acknowledge the support of the data providers and Trusted Research Environments that enabled secure access to the source datasets: SAIL Databank^22–24^ for BiW, University of Edinburgh DataStore and NHS Lothian ACCORD^13,45,46^ for Born in Scotland, SLaM Biomedical Research Centre (BRC) secure data environment, overseen by the Clinical Record Interactive Search (CRIS) Data Linkages service^17,47^ for Born in South London, Bradford Institute for Health Research^48^ (BIHR) for Born in Bradford cohorts, and the Birmingham Environment for Academic Research (BEAR) platform^18,29,49^ for CPRD.

