## Appendices for "Mother-infant linked UK electronic birth cohorts representing 17.5 million births harmonised to the OMOP common data model"

### Appendix A: Core data for birth cohorts

|  | **Dataset modules** | **Data** | **Details** | **Additional information** |
| --- | --- | --- | --- | --- |
| **Mother** | **Socio-demographics** | Week/Date of birth | Include time where available |  |
|  |  | Ethnicity |  |  |
|  |  | Deprivation Quintile | By nation at start of pregnancy | Assign by date and extract relative to event date for longitudinal data |
|  |  | Rural/urban | By nation at start of pregnancy | Assign by date and extract relative to event date for longitudinal data |
|  | **Pregnancy** | Date of antenatal visit |  |  |
|  |  | Pregancy start date | Estimated date of last menstrual period |  |
|  |  | Pregnancy end date | Date of delivery |  |
|  |  | Maternal smoking status |  | Linked to booking visit and later delivery dates |
|  |  | Maternal Height | in metres up to 2 decimal places |  |
|  |  | Maternal Weight | booking weight in kg up to 2 decimal places |  |
|  |  | Maternal BMI | kg/m^2^ to 2 decimal places |  |
|  |  | Existing comorbidities | Diagnostic codes leading to booking visit | Extract from primary and secondary care event codes before booking date |
|  |  | New events* | New events since booking visit | Extract from primary and secondary care event codes on/after booking date |
|  |  | Gravidity |  |  |
|  |  | Parity |  |  |
|  |  | Gestational age | At initial/booking antenatal visit | Derived from pregnancy start date and booking visit date |
|  | **Post-birth** | New events* | New events since delivery date | Extract from primary and secondary care event codes that first occur on/after delivery date. |
| **Infant/Child** | **Socio-demographics** | Week/Date of birth |  | Wales has week of birth - acquiring permission for date of birth |
|  |  | Sex | Sex at birth | Only male and female included; missing are excluded. |
|  |  | Ethnicity |  |  |
|  |  | Deprivation quintile | By locality at birth and school age | Assign by date and extract relative to event date for longitudinal data |
|  |  | Rural/urban residence | By locality at birth and school age | Assign by date and extract relative to event date for longitudinal data |
|  | **Birth** | Date of delivery |  |  |
|  |  | Gestational age | At birth | Derived from pregnancy start date and pregnancy end date. |
|  |  | Birth weight | in grams |  |
|  |  | New Events* | All primary and secondary care event codes from birth |  |
|  |  | Mode of delivery | E.g. c-section, forceps, vaginal, etc | Assigned to baby; else cannot assign different birth modes on multiple birth deliveries |
| **Events consist of code that represent diagnoses, procedures, prescriptions, devices and other health care administrative data.* | | | | |

### Appendix B: Metadata for raw data transformation of MIREDA birth cohorts core data

*Std = standard OMOP concept, Vocab. = Vocabulary, T = TRUE.*

| **Source  Vocab** | **Source  Field** | **Source  Value** | **Source  Term** | **Concept  ID** | **OMOP  Term** | **Class** | **Std** | **Valid** | **Domain** | **Vocab.** |
| --- | --- | --- | --- | --- | --- | --- | --- | --- | --- | --- |
| Non-Standard | GENDER | F | Female | 8532 | FEMALE | Gender | T | T | Gender | Gender |
| Non-Standard | GENDER | M | Male | 8507 | MALE | Gender | T | T | Gender | Gender |
| Non-Standard | BIRTH_OUTCOME | LIVEBIRTH | Live birth | 4092289 | Livebirth | Clinical Finding | T | T | Condition | SNOMED |
| Non-Standard | BIRTH_OUTCOME | STILLBIRTH | Stillbirth | 443213 | Stillbirth | Clinical Finding | T | T | Condition | SNOMED |
| Non-Standard | PREGNANCY_SINGLE | 0 | Multiple birth | 4163851 | Multiple birth | Clinical Finding | T | T | Condition | SNOMED |
| Non-Standard | PREGNANCY_SINGLE | 1 | Singleton birth | 1244607 | Singleton pregnancy | Clinical Finding | T | T | Observation | SNOMED |
| Non-Standard | GESTATIONAL_LENGTH_IN_DAY |  | Gestational age in days at birth | 4266763 | Fetal gestation at delivery - finding | Clinical Finding |  | T | Observation | SNOMED |
| Non-Standard | LABOUR_ONSET_GEST_WEEKS |  | Gastational age in weeks at labour onset | 4260747 | Length of gestation at birth | Observable Entity | T | T | Measurement | SNOMED |
| Non-Standard | BIRTH_WEIGHT |  | Birth weight (g) | 4264825 | Birth weight | Observable Entity | T | T | Measurement | SNOMED |
| Non-Standard | BIRTH_WEIGHT_CAT | ELBW | Extremely low birth weight less than 1,000g | 4173323 | Extremely low birth weight infant | Disorder | T | T | Condition | SNOMED |
| Non-Standard | BIRTH_WEIGHT_CAT | HBW | High birth weight 4,000 to 4,499g | 4079859 | High birth weight | Disorder | T | T | Condition | SNOMED |
| Non-Standard | BIRTH_WEIGHT_CAT | LBW | Low birth weight 1,500 to 2,499g | 4171115 | Low birth weight infant | Disorder | T | T | Condition | SNOMED |
| Non-Standard | BIRTH_WEIGHT_CAT | NBW | Normal birth weight 2,500 to 3,999g | 4080889 | Normal birth weight | Clinical Finding | T | T | Observation | SNOMED |
| Non-Standard | BIRTH_WEIGHT_CAT | VHBW | Very high birth weight 4,500g or greater | 434758 | Exceptionally large at birth | Disorder | T | T | Condition | SNOMED |
| Non-Standard | BIRTH_WEIGHT_CAT | VLBW | Very low birth weight 1,000 to 1,499g | 4079858 | Very low birth weight infant | Disorder | T | T | Condition | SNOMED |
| Non-Standard | PREGNANCY_MODE_DELIVERY | C-SECTION | C-section | 4015701 | Cesarean section | Procedure | T | T | Procedure | SNOMED |
| Non-Standard | PREGNANCY_MODE_DELIVERY | VAGINAL | Vaginal birth | 44784097 | Vaginal delivery of fetus | Procedure | T | T | Procedure | SNOMED |
| Non-Standard | BIRTH_MODE_CD | 1 | Spontaneous Vaginal Birth | 4071507 | Normal delivery procedure | Procedure | T | T | Procedure | SNOMED |
| Non-Standard | BIRTH_MODE_CD | 2 | Ventouse | 4189205 | Delivery by vacuum extraction | Procedure | T | T | Procedure | SNOMED |
| Non-Standard | BIRTH_MODE_CD | 3 | Forceps | 4114637 | Forceps delivery | Procedure | T | T | Procedure | SNOMED |
| Non-Standard | BIRTH_MODE_CD | 4 | Elective caesarean section . caesarean section before, or at onset of labour. | 4075182 | Elective cesarean section | Procedure | T | T | Procedure | SNOMED |
| Non-Standard | BIRTH_MODE_CD | 5 | Emergency caesarean section. | 4167089 | Emergency cesarean section | Procedure | T | T | Procedure | SNOMED |
| Non-Standard | PREGNANCY_CONCEPT | 4299535 | Pregancy | 4299535 | Pregnancy | Clinical Finding | T | T | Observation | SNOMED |
| Non-Standard | PREGNANCY_START_DATE |  | Date at the start of pregnancy/last menstrual period | 4072438 | Date of last menstrual period | Observable Entity | T | T | Measurement | SNOMED |
| Non-Standard | PREGNANCY_END_DATE |  | End of pregnancy date | 44816981 | Birth date of Fetus | Clinical Observation | T | T | Observation | LOINC |
| Non-Standard | PREGNANCY_HT |  | Height of mother in m | 607590 | Body height | Observable Entity | T | T | Measurement | SNOMED |
| Non-Standard | PREGNANCY_WT |  | Weight in kg at pregnancy booking. | 4178502 | Body weight measure | Observable Entity | T | T | Measurement | SNOMED |
| Non-Standard | PREGNANCY_BMI |  | Body mass index kg/m^2^ | 44807883 | Baseline body mass index | Observable Entity | T | T | Measurement | SNOMED |
| NHS Ethnicity Codes, Wales | RACE | A | WHITE - Any White Background, including: Welsh, English, Scottish, Northern Irish, Irish, British | 8527 | White | Race | T | T | Race | Race |
| NHS Ethnicity Codes, Wales | RACE | B | WHITE - Gypsy or Irish Traveller | 8527 | White | Race | T | T | Race | Race |
| NHS Ethnicity Codes, Wales | RACE | C | WHITE - White - Any other White background | 8527 | White | Race | T | T | Race | Race |
| NHS Ethnicity Codes, Wales | RACE | D | MIXED / MULTIPLE ETHNIC GROUP - White and Black Caribbean | 8527 | White | Race | T | T | Race | Race |
| NHS Ethnicity Codes, Wales | RACE | E | MIXED / MULTIPLE ETHNIC GROUP - White and Black African | 8527 | White | Race | T | T | Race | Race |
| NHS Ethnicity Codes, Wales | RACE | F | MIXED / MULTIPLE ETHNIC GROUP - White and Asian | 8527 | White | Race | T | T | Race | Race |
| NHS Ethnicity Codes, Wales | RACE | G | MIXED / MULTIPLE ETHNIC GROUP - Any other mixed background / multiple ethnic background | 8527 | White | Race | T | T | Race | Race |
| NHS Ethnicity Codes, Wales | RACE | H | ASIAN OR ASIAN BRITISH - Indian | 8515 | Asian | Race | T | T | Race | Race |
| NHS Ethnicity Codes, Wales | RACE | J | ASIAN OR ASIAN BRITISH - Pakistani | 8515 | Asian | Race | T | T | Race | Race |
| NHS Ethnicity Codes, Wales | RACE | K | ASIAN OR ASIAN BRITISH - Bangladeshi | 8515 | Asian | Race | T | T | Race | Race |
| NHS Ethnicity Codes, Wales | RACE | L | ASIAN OR ASIAN BRITISH - Any other Asian background | 8515 | Asian | Race | T | T | Race | Race |
| NHS Ethnicity Codes, Wales | RACE | M | BLACK OR BLACK BRITISH - Caribbean | 38003598 | Black | Race | T | T | Race | Race |
| NHS Ethnicity Codes, Wales | RACE | N | BLACK OR BLACK BRITISH - African | 38003598 | Black | Race | T | T | Race | Race |
| NHS Ethnicity Codes, Wales | RACE | P | BLACK OR BLACK BRITISH - Any other Black background | 38003598 | Black | Race | T | T | Race | Race |
| NHS Ethnicity Codes, Wales | RACE | R | ASIAN OR ASIAN BRITISH - Chinese | 8515 | Asian | Race | T | T | Race | Race |
| NHS Ethnicity Codes, Wales | RACE | A | WHITE - Any White Background, including: Welsh, English, Scottish, Northern Irish, Irish, British | 3959326 | White | Race |  | T | Observation | CO-CONNECT |
| NHS Ethnicity Codes, Wales | RACE | B | WHITE - Gypsy or Irish Traveller | 3959326 | White | Race |  | T | Observation | CO-CONNECT |
| NHS Ethnicity Codes, Wales | RACE | C | WHITE - White - Any other White background | 3959326 | White | Race |  | T | Observation | CO-CONNECT |
| NHS Ethnicity Codes, Wales | RACE | D | MIXED / MULTIPLE ETHNIC GROUP - White and Black Caribbean | 3959338 | Mixed Ethnic background | Race |  | T | Observation | CO-CONNECT |
| NHS Ethnicity Codes, Wales | RACE | E | MIXED / MULTIPLE ETHNIC GROUP - White and Black African | 3959338 | Mixed Ethnic background | Race |  | T | Observation | CO-CONNECT |
| NHS Ethnicity Codes, Wales | RACE | F | MIXED / MULTIPLE ETHNIC GROUP - White and Asian | 3959338 | Mixed Ethnic background | Race |  | T | Observation | CO-CONNECT |
| NHS Ethnicity Codes, Wales | RACE | G | MIXED / MULTIPLE ETHNIC GROUP - Any other mixed background / multiple ethnic background | 3959338 | Mixed Ethnic background | Race |  | T | Observation | CO-CONNECT |
| NHS Ethnicity Codes, Wales | RACE | H | ASIAN OR ASIAN BRITISH - Indian | 3959336 | Asian | Race |  | T | Observation | CO-CONNECT |
| NHS Ethnicity Codes, Wales | RACE | J | ASIAN OR ASIAN BRITISH - Pakistani | 3959336 | Asian | Race |  | T | Observation | CO-CONNECT |
| NHS Ethnicity Codes, Wales | RACE | K | ASIAN OR ASIAN BRITISH - Bangladeshi | 3959336 | Asian | Race |  | T | Observation | CO-CONNECT |
| NHS Ethnicity Codes, Wales | RACE | L | ASIAN OR ASIAN BRITISH - Any other Asian background | 3959336 | Asian | Race |  | T | Observation | CO-CONNECT |
| NHS Ethnicity Codes, Wales | RACE | M | BLACK OR BLACK BRITISH - Caribbean | 3959330 | Black | Race |  | T | Observation | CO-CONNECT |
| NHS Ethnicity Codes, Wales | RACE | N | BLACK OR BLACK BRITISH - African | 3959330 | Black | Race |  | T | Observation | CO-CONNECT |
| NHS Ethnicity Codes, Wales | RACE | P | BLACK OR BLACK BRITISH - Any other Black background | 3959330 | Black | Race |  | T | Observation | CO-CONNECT |
| NHS Ethnicity Codes, Wales | RACE | R | ASIAN OR ASIAN BRITISH - Chinese | 3959336 | Asian | Race |  | T | Observation | CO-CONNECT |
| NHS Ethnicity Codes, Wales | RACE | T | OTHER ETHNIC GROUPS - Arab | 3959547 | Other ethnic group | Race |  | T | Observation | CO-CONNECT |
| NHS Ethnicity Codes, Wales | RACE | D | MIXED / MULTIPLE ETHNIC GROUP - White and Black Caribbean | 3959542 | White and Black Caribbean | Race |  | T | Observation | CO-CONNECT |
| NHS Ethnicity Codes, Wales | RACE | E | MIXED / MULTIPLE ETHNIC GROUP - White and Black African | 3959543 | White and Black African | Race |  | T | Observation | CO-CONNECT |
| NHS Ethnicity Codes, Wales | RACE | F | MIXED / MULTIPLE ETHNIC GROUP - White and Asian | 3959544 | White and Asian | Race |  | T | Observation | CO-CONNECT |
| NHS Ethnicity Codes, Wales | RACE | G | MIXED / MULTIPLE ETHNIC GROUP - Any other mixed background / multiple ethnic background | 3959545 | Any other mixed background | Race |  | T | Observation | CO-CONNECT |
| NHS Ethnicity Codes, Wales | RACE | H | ASIAN OR ASIAN BRITISH - Indian | 3959333 | Asian Indian | Race |  | T | Observation | CO-CONNECT |
| NHS Ethnicity Codes, Wales | RACE | J | ASIAN OR ASIAN BRITISH - Pakistani | 3959331 | Asian Pakistani | Race |  | T | Observation | CO-CONNECT |
| NHS Ethnicity Codes, Wales | RACE | K | ASIAN OR ASIAN BRITISH - Bangladeshi | 3959332 | Asian Bangladeshi | Race |  | T | Observation | CO-CONNECT |
| NHS Ethnicity Codes, Wales | RACE | L | ASIAN OR ASIAN BRITISH - Any other Asian background | 3959335 | Any other asian background | Race |  | T | Observation | CO-CONNECT |
| NHS Ethnicity Codes, Wales | RACE | M | BLACK OR BLACK BRITISH - Caribbean | 3959328 | Black Caribbean | Race |  | T | Observation | CO-CONNECT |
| NHS Ethnicity Codes, Wales | RACE | N | BLACK OR BLACK BRITISH - African | 3959327 | Black African | Race |  | T | Observation | CO-CONNECT |
| NHS Ethnicity Codes, Wales | RACE | P | BLACK OR BLACK BRITISH - Any other Black background | 3959329 | Any other black background | Race |  | T | Observation | CO-CONNECT |
| NHS Ethnicity Codes, Wales | RACE | R | ASIAN OR ASIAN BRITISH - Chinese | 3959334 | Asian Chinese | Race |  | T | Observation | CO-CONNECT |
| NHS Ethnicity Codes, Wales | RACE | S | OTHER ETHNIC GROUPS - Any other ethnic group | 3959547 | Other ethnic group | Race |  | T | Observation | CO-CONNECT |
| NHS Ethnicity Codes, Wales | RACE | T | OTHER ETHNIC GROUPS - Arab | 359546 | Arab | Race |  | T | Observation | CO-CONNECT |
| Non-Standard | WIMD |  | Welsh Index of Multiple Deprivation (WIMD) Quintiles (1 = least deprived) | 35812898 | Index of Multiple Deprivation (Wales) | Variable |  | T | Observation | UK Biobank |
| Non-Standard | MAT_IA_SMOKING_CD | 1 | Smoker CO validated | 4298794 | Smoker | Clinical Finding | T | T | Observation | SNOMED |
| Non-Standard | MAT_IA_SMOKING_CD | 2 | Smoker Self Reported | 4298794 | Smoker | Clinical Finding | T | T | Observation | SNOMED |
| Non-Standard | MAT_IA_SMOKING_CD | 3 | Non Smoker CO validated | 4222303 | Non-Smoker | Clinical Finding | T | T | Observation | SNOMED |
| Non-Standard | MAT_IA_SMOKING_CD | 4 | Non Smoker Self Reported | 4222303 | Non-Smoker | Clinical Finding | T | T | Observation | SNOMED |
| Non-Standard | MAT_SMOKING_LABR_CD | 1 | Smoker CO validated | 4298794 | Smoker | Clinical Finding | T | T | Observation | SNOMED |
| Non-Standard | MAT_SMOKING_LABR_CD | 2 | Smoker Self Reported | 4298794 | Smoker | Clinical Finding | T | T | Observation | SNOMED |
| Non-Standard | MAT_SMOKING_LABR_CD | 3 | Non Smoker CO validated | 4222303 | Non-Smoker | Clinical Finding | T | T | Observation | SNOMED |
| Non-Standard | MAT_SMOKING_LABR_CD | 4 | Non Smoker Self Reported | 4222303 | Non-Smoker | Clinical Finding | T | T | Observation | SNOMED |
| ICD10 | EVENT_CD | O24.4 | Gestational diabetes | 4024659 | Gestational diabetes mellitus | Disorder | T | T | Condition | SNOMED |
| Read | EVENT_CD | L180100 | Gestational diabetes | 194700 | Diabetes mellitus in mother complicating pregnancy, childbirth AND/OR puerperium | Disorder | T | T | Condition | SNOMED |
| Read | EVENT_CD | L180300 | Gestational diabetes | 194700 | Diabetes mellitus in mother complicating pregnancy, childbirth AND/OR puerperium | Disorder | T | T | Condition | SNOMED |
| Read | EVENT_CD | L180800 | Gestational diabetes | 4024659 | Gestational diabetes mellitus | Disorder | T | T | Condition | SNOMED |
| Read | EVENT_CD | L180900 | Gestational diabetes | 4024659 | Gestational diabetes mellitus | Disorder | T | T | Condition | SNOMED |
| ICD10 | EVENT_CD | O13 | Hypertension | 4167493 | Pregnancy-induced hypertension | Disorder | T | T | Condition | SNOMED |
| Read | EVENT_CD | L123.00 | Hypertension | 441922 | Transient hypertension of pregnancy | Disorder | T | T | Condition | SNOMED |
| Read | EVENT_CD | L123000 | Hypertension | 441922 | Transient hypertension of pregnancy | Disorder | T | T | Condition | SNOMED |
| Read | EVENT_CD | L123100 | Hypertension | 441922 | Transient hypertension of pregnancy | Disorder | T | T | Condition | SNOMED |
| Read | EVENT_CD | L123200 | Hypertension | 4062906 | Transient hypertension of pregnancy with postnatal complication | Disorder | T | T | Condition | SNOMED |
| Read | EVENT_CD | L123300 | Hypertension | 441922 | Transient hypertension of pregnancy | Disorder | T | T | Condition | SNOMED |
| Read | EVENT_CD | L123400 | Hypertension | 4062906 | Transient hypertension of pregnancy with postnatal complication | Disorder | T | T | Condition | SNOMED |
| Read | EVENT_CD | L123500 | Hypertension | 4167493 | Pregnancy-induced hypertension | Disorder | T | T | Condition | SNOMED |
| Read | EVENT_CD | L123600 | Hypertension | 441922 | Transient hypertension of pregnancy | Disorder | T | T | Condition | SNOMED |
| Read | EVENT_CD | L123z00 | Hypertension | 441922 | Transient hypertension of pregnancy | Disorder | T | T | Condition | SNOMED |
| Read | EVENT_CD | L128.00 | Hypertension | 321074 | Pre-existing hypertension complicating pregnancy, childbirth and puerperium | Disorder | T | T | Condition | SNOMED |
| Read | EVENT_CD | L128000 | Hypertension | 4062552 | Pre-existing hypertensive heart disease complicating pregnancy, childbirth and the puerperium | Disorder | T | T | Condition | SNOMED |
| Read | EVENT_CD | L128100 | Hypertension | 4057978 | Pre-existing hypertensive heart and renal disease complicating pregnancy, childbirth and the puerperium | Disorder | T | T | Condition | SNOMED |
| Read | EVENT_CD | L128200 | Hypertension | 4057979 | Pre-existing secondary hypertension complicating pregnancy, childbirth and puerperium | Disorder | T | T | Condition | SNOMED |
| Read | EVENT_CD | L129.00 | Hypertension | 4034094 | Moderate proteinuric hypertension of pregnancy | Disorder | T | T | Condition | SNOMED |
| ICD10 | EVENT_CD | O11 | Pre-eclampsia | 4283352 | Pre-eclampsia added to pre-existing hypertension | Disorder | T | T | Condition | SNOMED |
| ICD10 | EVENT_CD | O14 | Pre-eclampsia | 439393 | Pre-eclampsia | Disorder | T | T | Condition | SNOMED |
| ICD10 | EVENT_CD | O14.0 | Pre-eclampsia | 439393 | Pre-eclampsia | Disorder | T | T | Condition | SNOMED |
| ICD10 | EVENT_CD | O14.1 | Pre-eclampsia | 433536 | Severe pre-eclampsia | Disorder | T | T | Condition | SNOMED |
| ICD10 | EVENT_CD | O14.2 | Pre-eclampsia | 4316372 | HELLP syndrome | Disorder | T | T | Condition | SNOMED |
| ICD10 | EVENT_CD | O14.9 | Pre-eclampsia | 439393 | Pre-eclampsia | Disorder | T | T | Condition | SNOMED |
| Read | EVENT_CD | L124.00 | Pre-eclampsia | 439393 | Pre-eclampsia | Disorder | T | T | Condition | SNOMED |
| Read | EVENT_CD | L124000 | Pre-eclampsia | 439393 | Pre-eclampsia | Disorder | T | T | Condition | SNOMED |
| Read | EVENT_CD | L124100 | Pre-eclampsia | 439393 | Pre-eclampsia | Disorder | T | T | Condition | SNOMED |
| Read | EVENT_CD | L124200 | Pre-eclampsia | 439393 | Pre-eclampsia | Disorder | T | T | Condition | SNOMED |
| Read | EVENT_CD | L124300 | Pre-eclampsia | 439393 | Pre-eclampsia | Disorder | T | T | Condition | SNOMED |
| Read | EVENT_CD | L124400 | Pre-eclampsia | 439393 | Pre-eclampsia | Disorder | T | T | Condition | SNOMED |
| Read | EVENT_CD | L124500 | Pre-eclampsia | 314090 | Mild pre-eclampsia | Disorder | T | T | Condition | SNOMED |
| Read | EVENT_CD | L124600 | Pre-eclampsia | 439393 | Pre-eclampsia | Disorder | T | T | Condition | SNOMED |
| Read | EVENT_CD | L124z00 | Pre-eclampsia | 439393 | Pre-eclampsia | Disorder | T | T | Condition | SNOMED |
| Read | EVENT_CD | L125.00 | Pre-eclampsia | 433536 | Severe pre-eclampsia | Disorder | T | T | Condition | SNOMED |
| Read | EVENT_CD | L125000 | Pre-eclampsia | 433536 | Severe pre-eclampsia | Disorder | T | T | Condition | SNOMED |
| Read | EVENT_CD | L125100 | Pre-eclampsia | 433536 | Severe pre-eclampsia | Disorder | T | T | Condition | SNOMED |
| Read | EVENT_CD | L125200 | Pre-eclampsia | 433536 | Severe pre-eclampsia | Disorder | T | T | Condition | SNOMED |
| Read | EVENT_CD | L125400 | Pre-eclampsia | 4057976 | Severe pre-eclampsia with postnatal complication | Disorder | T | T | Condition | SNOMED |
| Read | EVENT_CD | L125z00 | Pre-eclampsia | 433536 | Severe pre-eclampsia | Disorder | T | T | Condition | SNOMED |
| Read | EVENT_CD | L12A.00 | Pre-eclampsia | 4316372 | HELLP syndrome | Disorder | T | T | Condition | SNOMED |
| Read | EVENT_CD | L12B.00 | Pre-eclampsia | 439393 | Pre-eclampsia | Disorder | T | T | Condition | SNOMED |
| Read | EVENT_CD | L12z000 | Pre-eclampsia | 4167493 | Pregnancy-induced hypertension | Disorder | T | T | Condition | SNOMED |
| Read | EVENT_CD | L12z100 | Pre-eclampsia | 4167493 | Pregnancy-induced hypertension | Disorder | T | T | Condition | SNOMED |
| Read | EVENT_CD | L12z200 | Pre-eclampsia | 4167493 | Pregnancy-induced hypertension | Disorder | T | T | Condition | SNOMED |
| Read | EVENT_CD | L12z300 | Pre-eclampsia | 4167493 | Pregnancy-induced hypertension | Disorder | T | T | Condition | SNOMED |
| Read | EVENT_CD | L12z400 | Pre-eclampsia | 4167493 | Pregnancy-induced hypertension | Disorder | T | T | Condition | SNOMED |
| Read | EVENT_CD | L12zz00 | Pre-eclampsia | 4167493 | Pregnancy-induced hypertension | Disorder | T | T | Condition | SNOMED |
| Read | EVENT_CD | L127.00 | Pre-/Eclampsia | 141084 | Pre-eclampsia or eclampsia with pre-existing hypertension | Disorder | T | T | Condition | SNOMED |
| Read | EVENT_CD | L127000 | Pre-/Eclampsia | 141084 | Pre-eclampsia or eclampsia with pre-existing hypertension | Disorder | T | T | Condition | SNOMED |
| Read | EVENT_CD | L127400 | Pre-/Eclampsia | 4062550 | Pre-eclampsia or eclampsia with pre-existing hypertension with postnatal complication | Disorder | T | T | Condition | SNOMED |
| Read | EVENT_CD | L127z00 | Pre-/Eclampsia | 141084 | Pre-eclampsia or eclampsia with pre-existing hypertension | Disorder | T | T | Condition | SNOMED |
| ICD10 | EVENT_CD | O15 | Eclampsia | 443700 | Eclampsia | Disorder | T | T | Condition | SNOMED |
| ICD10 | EVENT_CD | O15.0 | Eclampsia | 137613 | Eclampsia in pregnancy | Disorder | T | T | Condition | SNOMED |
| ICD10 | EVENT_CD | O15.1 | Eclampsia | 4034096 | Eclampsia in labor | Disorder | T | T | Condition | SNOMED |
| ICD10 | EVENT_CD | O15.2 | Eclampsia | 4116344 | Eclampsia in puerperium | Disorder | T | T | Condition | SNOMED |
| ICD10 | EVENT_CD | O15.9 | Eclampsia | 443700 | Eclampsia | Disorder | T | T | Condition | SNOMED |
| Read | EVENT_CD | L126.00 | Eclampsia | 443700 | Eclampsia | Disorder | T | T | Condition | SNOMED |
| Read | EVENT_CD | L126000 | Eclampsia | 443700 | Eclampsia | Disorder | T | T | Condition | SNOMED |
| Read | EVENT_CD | L126100 | Eclampsia | 443700 | Eclampsia | Disorder | T | T | Condition | SNOMED |
| Read | EVENT_CD | L126200 | Eclampsia | 4058536 | Eclampsia with postnatal complication | Disorder | T | T | Condition | SNOMED |
| Read | EVENT_CD | L126300 | Eclampsia | 137613 | Eclampsia in pregnancy | Disorder | T | T | Condition | SNOMED |
| Read | EVENT_CD | L126400 | Eclampsia | 4058536 | Eclampsia with postnatal complication | Disorder | T | T | Condition | SNOMED |
| Read | EVENT_CD | L126500 | Eclampsia | 137613 | Eclampsia in pregnancy | Disorder | T | T | Condition | SNOMED |
| Read | EVENT_CD | L126600 | Eclampsia | 4034096 | Eclampsia in labor | Disorder | T | T | Condition | SNOMED |
| Read | EVENT_CD | L126z00 | Eclampsia | 443700 | Eclampsia | Disorder | T | T | Condition | SNOMED |
| ICD10 | EVENT_CD | D50 | Anaemias in pregnancy | 436659 | Iron deficiency anemia | Disorder | T | T | Condition | SNOMED |
| ICD10 | EVENT_CD | D50.0 | Anaemias in pregnancy | 37119138 | Iron deficiency anemia due to blood loss | Disorder | T | T | Condition | SNOMED |
| ICD10 | EVENT_CD | D50.1 | Anaemias in pregnancy | 4198185 | Plummer-Vinson syndrome | Disorder | T | T | Condition | SNOMED |
| ICD10 | EVENT_CD | D50.8 | Anaemias in pregnancy | 436659 | Iron deficiency anemia | Disorder | T | T | Condition | SNOMED |
| ICD10 | EVENT_CD | D50.9 | Anaemias in pregnancy | 436659 | Iron deficiency anemia | Disorder | T | T | Condition | SNOMED |
| ICD10 | EVENT_CD | D51 | Anaemias in pregnancy | 432588 | Megaloblastic anemia due to vitamin B-12 deficiency | Disorder | T | T | Condition | SNOMED |
| ICD10 | EVENT_CD | D51.0 | Anaemias in pregnancy | 432295 | Pernicious anemia | Disorder | T | T | Condition | SNOMED |
| ICD10 | EVENT_CD | D51.1 | Anaemias in pregnancy | 4096927 | Megaloblastic anemia due to vitamin B-12 malabsorption with proteinuria | Disorder | T | T | Condition | SNOMED |
| ICD10 | EVENT_CD | D51.2 | Anaemias in pregnancy | 4029726 | Transcobalamin II deficiency | Disorder | T | T | Condition | SNOMED |
| ICD10 | EVENT_CD | D51.3 | Anaemias in pregnancy | 4147491 | Vitamin B12 deficiency anemia due to dietary causes | Disorder | T | T | Condition | SNOMED |
| ICD10 | EVENT_CD | D51.8 | Anaemias in pregnancy | 432588 | Megaloblastic anemia due to vitamin B-12 deficiency | Disorder | T | T | Condition | SNOMED |
| ICD10 | EVENT_CD | D51.9 | Anaemias in pregnancy | 432588 | Megaloblastic anemia due to vitamin B-12 deficiency | Disorder | T | T | Condition | SNOMED |
| ICD10 | EVENT_CD | D52 | Anaemias in pregnancy | 440977 | Megaloblastic anemia due to folate deficiency | Disorder | T | T | Condition | SNOMED |
| ICD10 | EVENT_CD | D52.0 | Anaemias in pregnancy | 4143351 | Folate deficiency anemia due to dietary causes | Disorder | T | T | Condition | SNOMED |
| ICD10 | EVENT_CD | D52.1 | Anaemias in pregnancy | 4143351 | Folate deficiency anemia due to dietary causes | Disorder | T | T | Condition | SNOMED |
| ICD10 | EVENT_CD | D52.8 | Anaemias in pregnancy | 440977 | Megaloblastic anemia due to folate deficiency | Disorder | T | T | Condition | SNOMED |
| ICD10 | EVENT_CD | D52.9 | Anaemias in pregnancy | 440977 | Megaloblastic anemia due to folate deficiency | Disorder | T | T | Condition | SNOMED |
| ICD10 | EVENT_CD | O99.0 | Anaemias in pregnancy | 434701 | Anemia in mother complicating pregnancy, childbirth AND/OR puerperium | Disorder | T | T | Condition | SNOMED |
| Read | EVENT_CD | D001.00 | Anaemias in pregnancy | 4100985 | Iron deficiency anemia due to dietary causes | Disorder | T | T | Condition | SNOMED |
| Read | EVENT_CD | D011000 | Anaemias in pregnancy | 4147491 | Vitamin B12 deficiency anemia due to dietary causes | Disorder | T | T | Condition | SNOMED |
| Read | EVENT_CD | D011X00 | Anaemias in pregnancy | 432588 | Megaloblastic anemia due to vitamin B-12 deficiency | Disorder | T | T | Condition | SNOMED |
| Read | EVENT_CD | D011z00 | Anaemias in pregnancy | 432588 | Megaloblastic anemia due to vitamin B-12 deficiency | Disorder | T | T | Condition | SNOMED |
| Read | EVENT_CD | D012100 | Anaemias in pregnancy | 4143351 | Folate deficiency anemia due to dietary causes | Disorder | T | T | Condition | SNOMED |
| Read | EVENT_CD | D012200 | Anaemias in pregnancy | 4100987 | Folate deficiency anemia, drug-induced | Disorder | T | T | Condition | SNOMED |
| Read | EVENT_CD | D012300 | Anaemias in pregnancy | 4098008 | Folate deficiency anemia due to malabsorption | Disorder | T | T | Condition | SNOMED |
| Read | EVENT_CD | D012400 | Anaemias in pregnancy | 4098009 | Folate deficiency anemia due to liver disorders | Disorder | T | T | Condition | SNOMED |
| Read | EVENT_CD | D012500 | Anaemias in pregnancy | 4308125 | Macrocytic anemia | Disorder | T | T | Condition | SNOMED |
| Read | EVENT_CD | D012z00 | Anaemias in pregnancy | 440977 | Megaloblastic anemia due to folate deficiency | Disorder | T | T | Condition | SNOMED |
| Read | EVENT_CD | D013000 | Anaemias in pregnancy | 4101458 | Combined B12 and folate deficiency anemia | Disorder | T | T | Condition | SNOMED |
| Read | EVENT_CD | L182100 | Anaemias in pregnancy | 4099889 | Anemia of pregnancy | Disorder | T | T | Condition | SNOMED |
| Read | EVENT_CD | L182300 | Anaemias in pregnancy | 4099889 | Anemia of pregnancy | Disorder | T | T | Condition | SNOMED |
| Read | EVENT_CD | L182500 | Anaemias in pregnancy | 4058246 | Iron deficiency anemia of pregnancy | Disorder | T | T | Condition | SNOMED |
| ICD10 | EVENT_CD | F31.3 | Depression in pregnancy | 439254 | Bipolar affective disorder, current episode depression | Disorder | T | T | Condition | SNOMED |
| ICD10 | EVENT_CD | F31.4 | Depression in pregnancy | 439254 | Bipolar affective disorder, current episode depression | Disorder | T | T | Condition | SNOMED |
| ICD10 | EVENT_CD | F31.5 | Depression in pregnancy | 35622934 | Psychosis and severe depression co-occurrent and due to bipolar affective disorder | Disorder | T | T | Condition | SNOMED |
| ICD10 | EVENT_CD | F31.6 | Depression in pregnancy | 435226 | Bipolar affective disorder, current episode mixed | Disorder | T | T | Condition | SNOMED |
| ICD10 | EVENT_CD | F32 | Depression in pregnancy | 440383 | Depressive disorder | Disorder | T | T | Condition | SNOMED |
| ICD10 | EVENT_CD | F32.0 | Depression in pregnancy | 4195572 | Mild major depression, single episode | Disorder | T | T | Condition | SNOMED |
| ICD10 | EVENT_CD | F32.1 | Depression in pregnancy | 4049623 | Moderate major depression, single episode | Disorder | T | T | Condition | SNOMED |
| ICD10 | EVENT_CD | F32.2 | Depression in pregnancy | 441534 | Severe major depression, single episode, without psychotic features | Disorder | T | T | Condition | SNOMED |
| ICD10 | EVENT_CD | F32.3 | Depression in pregnancy | 438406 | Severe major depression, single episode, with psychotic features | Disorder | T | T | Condition | SNOMED |
| ICD10 | EVENT_CD | F32.8 | Depression in pregnancy | 440383 | Depressive disorder | Disorder | T | T | Condition | SNOMED |
| ICD10 | EVENT_CD | F32.9 | Depression in pregnancy | 3656234 | Depressive episode | Clinical Finding | T | T | Observation | SNOMED |
| ICD10 | EVENT_CD | F33 | Depression in pregnancy | 4098302 | Recurrent depression | Disorder | T | T | Condition | SNOMED |
| ICD10 | EVENT_CD | F33.0 | Depression in pregnancy | 4228802 | Mild recurrent major depression | Disorder | T | T | Condition | SNOMED |
| ICD10 | EVENT_CD | F33.1 | Depression in pregnancy | 4077577 | Moderate recurrent major depression | Disorder | T | T | Condition | SNOMED |
| ICD10 | EVENT_CD | F33.2 | Depression in pregnancy | 435220 | Severe recurrent major depression without psychotic features | Disorder | T | T | Condition | SNOMED |
| ICD10 | EVENT_CD | F33.3 | Depression in pregnancy | 434911 | Recurrent major depressive episodes, severe, with psychosis | Disorder | T | T | Condition | SNOMED |
| ICD10 | EVENT_CD | F33.8 | Depression in pregnancy | 4098302 | Recurrent depression | Disorder | T | T | Condition | SNOMED |
| ICD10 | EVENT_CD | F33.9 | Depression in pregnancy | 4282316 | Recurrent major depression | Disorder | T | T | Condition | SNOMED |
| Read | EVENT_CD | E113.00 | Depression in pregnancy | 432285 | Recurrent major depressive episodes | Disorder | T | T | Condition | SNOMED |
| Read | EVENT_CD | E113000 | Depression in pregnancy | 432285 | Recurrent major depressive episodes | Disorder | T | T | Condition | SNOMED |
| Read | EVENT_CD | E113100 | Depression in pregnancy | 438998 | Recurrent major depressive episodes, mild | Disorder | T | T | Condition | SNOMED |
| Read | EVENT_CD | E113200 | Depression in pregnancy | 432883 | Recurrent major depressive episodes, moderate | Disorder | T | T | Condition | SNOMED |
| Read | EVENT_CD | E113300 | Depression in pregnancy | 44805542 | Recurrent major depressive episodes, severe | Disorder | T | T | Condition | SNOMED |
| Read | EVENT_CD | E113400 | Depression in pregnancy | 434911 | Recurrent major depressive episodes, severe, with psychosis | Disorder | T | T | Condition | SNOMED |
| Read | EVENT_CD | E113500 | Depression in pregnancy | 44805549 | Recurrent major depressive episodes, in partial remission | Disorder | T | T | Condition | SNOMED |
| Read | EVENT_CD | E113600 | Depression in pregnancy | 4263748 | Recurrent major depression in full remission | Disorder | T | T | Condition | SNOMED |
| Read | EVENT_CD | E113700 | Depression in pregnancy | 4098302 | Recurrent depression | Disorder | T | T | Condition | SNOMED |
| Read | EVENT_CD | E113z00 | Depression in pregnancy | 432285 | Recurrent major depressive episodes | Disorder | T | T | Condition | SNOMED |
| Read | EVENT_CD | E115.00 | Depression in pregnancy | 439254 | Bipolar affective disorder, current episode depression | Disorder | T | T | Condition | SNOMED |
| Read | EVENT_CD | E115000 | Depression in pregnancy | 439254 | Bipolar affective disorder, current episode depression | Disorder | T | T | Condition | SNOMED |
| Read | EVENT_CD | E115100 | Depression in pregnancy | 439253 | Bipolar affective disorder, currently depressed, mild | Disorder | T | T | Condition | SNOMED |
| Read | EVENT_CD | E115200 | Depression in pregnancy | 437528 | Bipolar affective disorder, currently depressed, moderate | Disorder | T | T | Condition | SNOMED |
| Read | EVENT_CD | E115300 | Depression in pregnancy | 442570 | Severe depressed bipolar I disorder without psychotic features | Disorder | T | T | Condition | SNOMED |
| Read | EVENT_CD | E115400 | Depression in pregnancy | 35622934 | Psychosis and severe depression co-occurrent and due to bipolar affective disorder | Disorder | T | T | Condition | SNOMED |
| Read | EVENT_CD | E115500 | Depression in pregnancy | 4177651 | Depressed bipolar I disorder in partial remission | Disorder | T | T | Condition | SNOMED |
| Read | EVENT_CD | E115600 | Depression in pregnancy | 439251 | Bipolar affective disorder, currently depressed, in full remission | Disorder | T | T | Condition | SNOMED |
| Read | EVENT_CD | E115z00 | Depression in pregnancy | 439254 | Bipolar affective disorder, current episode depression | Disorder | T | T | Condition | SNOMED |
| Read | EVENT_CD | E116.00 | Depression in pregnancy | 439250 | Mixed bipolar affective disorder | Disorder | T | T | Condition | SNOMED |
| Read | EVENT_CD | E116000 | Depression in pregnancy | 439250 | Mixed bipolar affective disorder | Disorder | T | T | Condition | SNOMED |
| Read | EVENT_CD | E116100 | Depression in pregnancy | 439249 | Mixed bipolar affective disorder, mild | Disorder | T | T | Condition | SNOMED |
| Read | EVENT_CD | E116200 | Depression in pregnancy | 439248 | Mixed bipolar affective disorder, moderate | Disorder | T | T | Condition | SNOMED |
| Read | EVENT_CD | E116300 | Depression in pregnancy | 44805540 | Mixed bipolar affective disorder, severe | Disorder | T | T | Condition | SNOMED |
| Read | EVENT_CD | E116400 | Depression in pregnancy | 439246 | Mixed bipolar affective disorder, severe, with psychosis | Disorder | T | T | Condition | SNOMED |
| Read | EVENT_CD | E116500 | Depression in pregnancy | 44804961 | Mixed bipolar affective disorder, in partial remission | Disorder | T | T | Condition | SNOMED |
| Read | EVENT_CD | E116600 | Depression in pregnancy | 439245 | Mixed bipolar affective disorder, in full remission | Disorder | T | T | Condition | SNOMED |
| Read | EVENT_CD | E116z00 | Depression in pregnancy | 439250 | Mixed bipolar affective disorder | Disorder | T | T | Condition | SNOMED |
| Read | EVENT_CD | E117.00 | Depression in pregnancy | 436665 | Bipolar disorder | Disorder | T | T | Condition | SNOMED |
| Read | EVENT_CD | E117000 | Depression in pregnancy | 436665 | Bipolar disorder | Disorder | T | T | Condition | SNOMED |
| Read | EVENT_CD | E117100 | Depression in pregnancy | 4028027 | Mild bipolar disorder | Disorder | T | T | Condition | SNOMED |
| Read | EVENT_CD | E117200 | Depression in pregnancy | 4194222 | Moderate bipolar disorder | Disorder | T | T | Condition | SNOMED |
| Read | EVENT_CD | E117300 | Depression in pregnancy | 4200385 | Severe bipolar disorder without psychotic features | Disorder | T | T | Condition | SNOMED |
| Read | EVENT_CD | E117400 | Depression in pregnancy | 4195158 | Severe bipolar disorder with psychotic features | Disorder | T | T | Condition | SNOMED |
| Read | EVENT_CD | E117500 | Depression in pregnancy | 436072 | Bipolar disorder in partial remission | Disorder | T | T | Condition | SNOMED |
| Read | EVENT_CD | E117600 | Depression in pregnancy | 4220618 | Bipolar disorder in full remission | Disorder | T | T | Condition | SNOMED |
| Read | EVENT_CD | E117z00 | Depression in pregnancy | 436665 | Bipolar disorder | Disorder | T | T | Condition | SNOMED |
| Read | EVENT_CD | E118.00 | Depression in pregnancy | 4092239 | Seasonal affective disorder | Disorder | T | T | Condition | SNOMED |
| Read | EVENT_CD | E11y000 | Depression in pregnancy | 436665 | Bipolar disorder | Disorder | T | T | Condition | SNOMED |
| Read | EVENT_CD | E11y200 | Depression in pregnancy | 438727 | Atypical depressive disorder | Disorder | T | T | Condition | SNOMED |
| Read | EVENT_CD | E11yz00 | Depression in pregnancy | 436665 | Bipolar disorder | Disorder | T | T | Condition | SNOMED |
| Read | EVENT_CD | E11z200 | Depression in pregnancy | 4338029 | Masked depression | Disorder | T | T | Condition | SNOMED |
| Read | EVENT_CD | E200300 | Depression in pregnancy | 4338031 | Mixed anxiety and depressive disorder | Disorder | T | T | Condition | SNOMED |
| Read | EVENT_CD | E204.00 | Depression in pregnancy | 4314692 | Reactive depression (situational) | Disorder | T | T | Condition | SNOMED |
| Read | EVENT_CD | E2B1.00 | Depression in pregnancy | 4103574 | Chronic depression | Disorder | T | T | Condition | SNOMED |
| Non-Standard | PREV_PREGNANCY_PARITY | Multiparous | Multiparous | 4102166 | Multiparous | Clinical Finding | T | T | Condition | SNOMED |
| Non-Standard | PREV_PREGNANCY_PARITY | Nulliparous | Nulliparous | 4012561 | Nulliparous | Clinical Finding | T | T | Condition | SNOMED |
| Non-Standard | PREV_PREGNANCY_PARITY | Primiparous | Primiparous | 4030563 | Para 1 | Clinical Finding | T | T | Condition | SNOMED |
| Non-Standard | PREV_PREGNANCY_PARITY |  | Parity | 4041279 | Parity finding | Clinical Finding | T | T | Condition | SNOMED |
